# Perfusate metabolomics content and tubular transporters expression during kidney graft preservation by hypothermic machine perfusion

**DOI:** 10.1101/2021.09.27.21264167

**Authors:** Quentin Faucher, Hugo Alarcan, François-Ludovic Sauvage, Lionel Forestier, Elodie Miquelestorena-Standley, Lydie Nadal-Desbarats, Hélène Arnion, Jean-Christophe Venhard, Nicolas Brichart, Franck Bruyère, Pierre Marquet, Chantal Barin-Le Guellec

## Abstract

**Background:** Ischemia-related injury during the pre-implantation period impacts kidney graft outcome. Evaluating these lesions by a non-invasive approach before transplantation could help to understand the mechanisms of graft injury and identify potential biomarkers predictive of graft outcomes. This study aims to determine metabolomic content of graft perfusion fluids and its dependence on preservation time and to explore whether tubular transporters are possibly involved in the metabolomics variations observed.

**Methods:** Kidneys were stored on hypothermic perfusion machines. We evaluated the metabolomic profiles of perfusion fluids (n=35) using Liquid Chromatography coupled with tandem Mass Spectrometry and studied the transcriptional expression of tubular transporters on pre-implantation biopsies (n=26). We used univariate and multivariate analyses to assess the impact of perfusion time on these parameters and their relationship with graft outcome.

**Results:** Seventy-two metabolites were found in preservation fluids at the end of perfusion, of which 40% were already present in the native conservation solution. We observed an increase of 23 metabolites with longer perfusion time and a decrease for 8. The predictive model for time-dependent variation of metabolomics content showed good performance (R^2^= 76%, Q^2^= 54%, accuracy= 41%, permutations test significant). Perfusion time had no effect on the mRNA expression of transporters. We found no correlation between metabolomics and transporters expression. Neither the metabolomics profile nor the transporters expression were predictive of graft outcome.

**Conclusion:** Our results open the way for further studies, focusing on both intra- and extra-tissue metabolome, to investigate whether transporter alterations can explain the variations observed in pre-implantation period.

## Introduction

Kidney transplantation is the treatment of choice for patients suffering from end-stage renal diseases. According to the World Health Organization, the death rate from kidney diseases will continue to increase to reach 27 deaths per 100 000 people by 2060^1^, while the use of renal replacement therapy (i.e. dialysis or kidney transplantation) worldwide will reach ±5 million people by 2030^2^. With regard to this increase of renal diseases, the gap between the demand for organs and the limited group of donors will continue to widen. To overcome this issue, many centers are gradually accepting sub-optimal donors, including donation after circulatory death and Extended Criteria Donors (ECD). However, kidneys retrieved from such donors are more prone to Ischemia-Reperfusion Injury (IRI) and Delayed Graft Function (DGF) in the post-transplant period^3^. IRI is a multifactorial pathophysiological process incumbent to the transplantation procedure and a major cause of DGF, which in turn increases the risk of short- and long-term poor graft outcomes^4–7^. For these sub-optimal donors, Hypothermic Machine Perfusion (HMP), an *ex vivo* circulating, hypoxic environment, is recommended for kidney preservation, as compared to static cold storage (SCS), in order to reduce DGF rates and improve graft survival^8^. However, reliable tools are needed for the evaluation of graft quality during HMP preservation. Metabolomic analysis of the perfusion fluid provides the possibility to reveal potential biomarkers of graft quality or predictive of transplantation outcomes, but also to inform about the cellular mechanisms occurring during organ perfusion^9^. The few studies previously conducted on this topic showed a variation of the metabolomic content according to the perfusion time: mainly an increase of lactate and amino-acids and a decrease of glutathione^10–12^. These variations suggest metabolites are uptaken or released by the kidney during its preservation. Renal tubular membrane transporters (mainly of the SoLute Carrier (SLC) and ATP Binding-Cassette (ABC) families) play a major role in cell and tissue homeostasis owing to the bidirectional, transcellular exchanges they are involved in. Alteration of their activity, previously demonstrated (mainly for SLC) during ischemia or ischemia/reperfusion^13,14^, could be responsible for some metabolic variations observed during organ perfusion, but also have deleterious consequences for the graft outcomes^15^. Concerning the predictive value of the perfusion fluid metabolomic content, controversial results have been found and a recent review suggests that further studies are needed^16^.

The objectives of the present clinical study were to determine the metabolomic contents of perfusion fluids collected at the end of kidney graft perfusion on HMP and the transcriptional expression of renal tubular transporters on graft biopsies taken at the same time. We aimed to explore the relationships between ischemia duration, *ex situ* metabolites, transporters expression and graft outcome. To the best of our knowledge, this is the first evaluation of the potential impact of membrane transporter alterations on the variations of the perfusion fluid metabolomic contents observed during graft preservation by HMP.

## Materials and Methods

### A more detailed description of the materials and methods is available in the Supplementary Digital Content (SCD, “Material and Methods”)

As part of the clinical research project “RENALIFE” (ClinicalTrials.gov NCT03024229), 38 kidneys taken from ECD were included. Organs were stored on HMP LifePort^®^ Kidney Transporter 1.0 (Organ Recovery Systems) with KPS-1^®^ (Organ Recovery System) as preservation solution. HMP parameters (temperature, flow, resistance) were recorded during kidney conservation and a perfusate sample was collected at the end of perfusion (storage at -20°C after centrifugation: 3000g, 10min). Preimplantation biopsy (targeting the renal cortex) was performed and split in two, with one fragment embedded in paraffin after fixation for anatomopathological evaluation and the other stored at -20°C. Serum creatinine and clinical events were recorded up to 7 days and 3 months respectively. Immediate Graft Function (IGF) was characterized by a serum creatinine ≤ 250 µmol/L on day 7 without the need for dialysis and Delayed Graft Function (DGF) by the necessity of dialysis within the first 7 days. Mass spectrometry chromatographic analysis was performed using a LCMS-8060 tandem mass spectrometer (Shimadzu) and the package “Method Package for Cell Culture Profiling Ver.2” (Shimadzu). Infusion of pure substances was performed to add some molecules to the package. Perfusates were analyzed in duplicate and native KPS-1 was injected to determine its basal composition. A COBAS 6000 analyzer (Roche Diagnostics) was used to determine sodium, potassium, calcium, phosphate, chloride, bicarbonate, urea, creatinine, and glucose concentrations and add them to the previous metabolites.

Graft RNA was extracted from frozen pre-implantation biopsies. After quantification and integrity evaluation, retro-transcription was performed. Taqman Low Density Array (TLDA) cards were used to determine the transcriptional expression of 35 membrane tubular transporters, 3 aquaporins, 2 Na/K-ATPase sub-units (Supplementary table 1 for probe sets) and 4 housekeeping gene candidates (*NME4, CHFR, C16ORF62* and *NASP*) chosen according to the literature^17^. Undetermined or > 35 Ct values were replaced by 35. Transporters expression was analyzed by the comparative 2^-ΔCt^ method with ΔCt = Ct (target gene) – Ct (mean of the housekeeping genes finally retained)^18^. Then 2^-ΔCt^ were normalized by log2 transformation. Three housekeeping genes (*NME4, CHFR* and *C16ORF62*) were finally selected using Genorm^19^ and Normfinder^20^.

To explore the impact of ischemia duration on the perfusate metabolomics contents and tubular transporters expression, grafts were allocated to 3 different perfusion duration groups: < 12h, between 12 and 20h, and > 20h.

Statistical analysis involved univariate followed by multivariate analysis (Principal Component Analysis (PCA), Partial Least Squares discriminant analysis (PLS-DA), Partial Least Squares (PLS), Random Forest (RF), in accordance with standard approaches for metabolomic analyzes^21^). The MetaboAnalyst 5.0 computational platform (www.metaboanalyst.ca/faces/home.xhtml) was used for all the analyses except for PLS which was performed using the MixOmics package (version 1.6.3) in R (version 4.0.2).

## Results

### 1. Study population

Thirty-eight donor-recipients couples were included (Table 1). All donors were brain dead and ECD. The median perfusion and cold ischemia time was 831.5 and 1020 min, respectively.

**Table 1:**
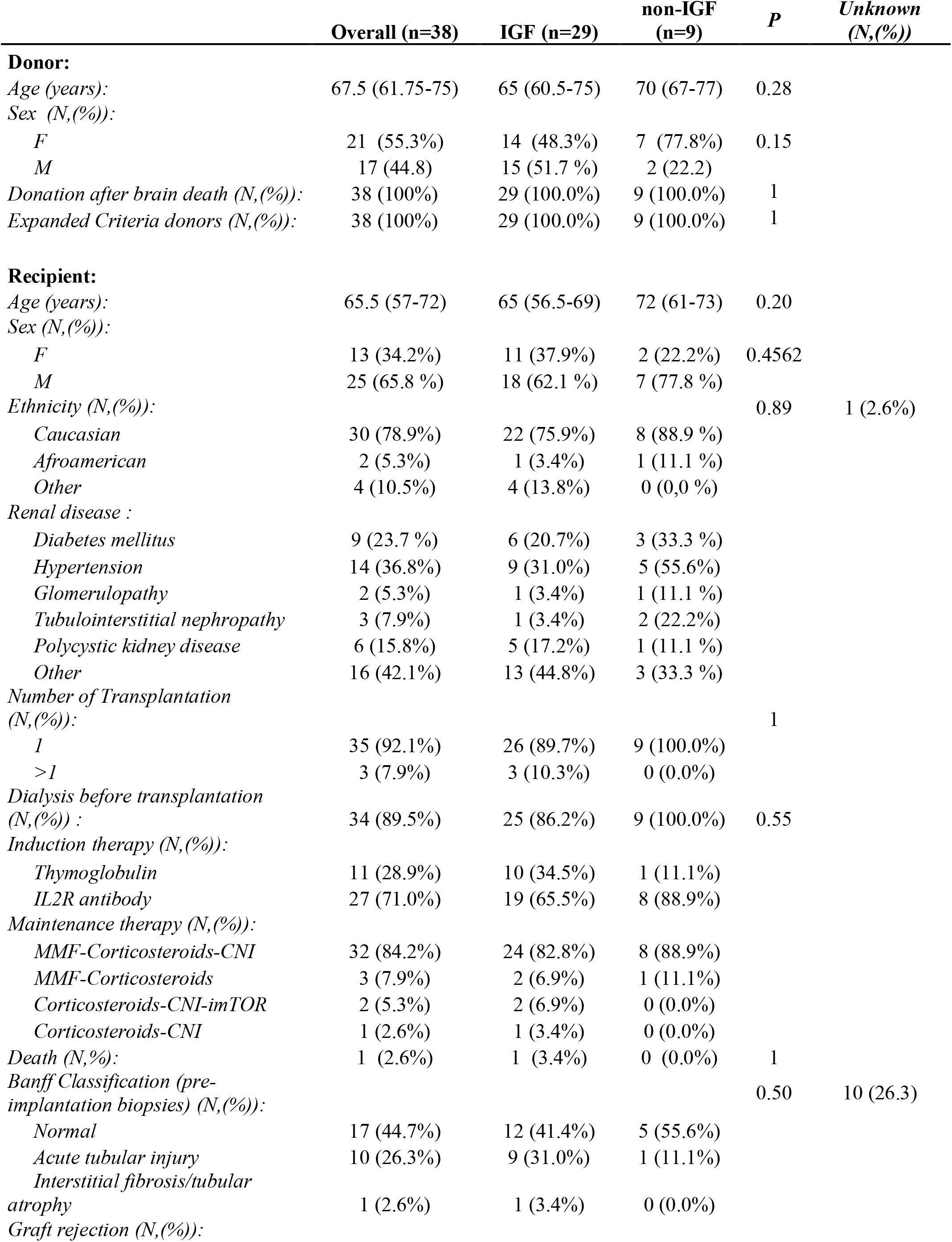
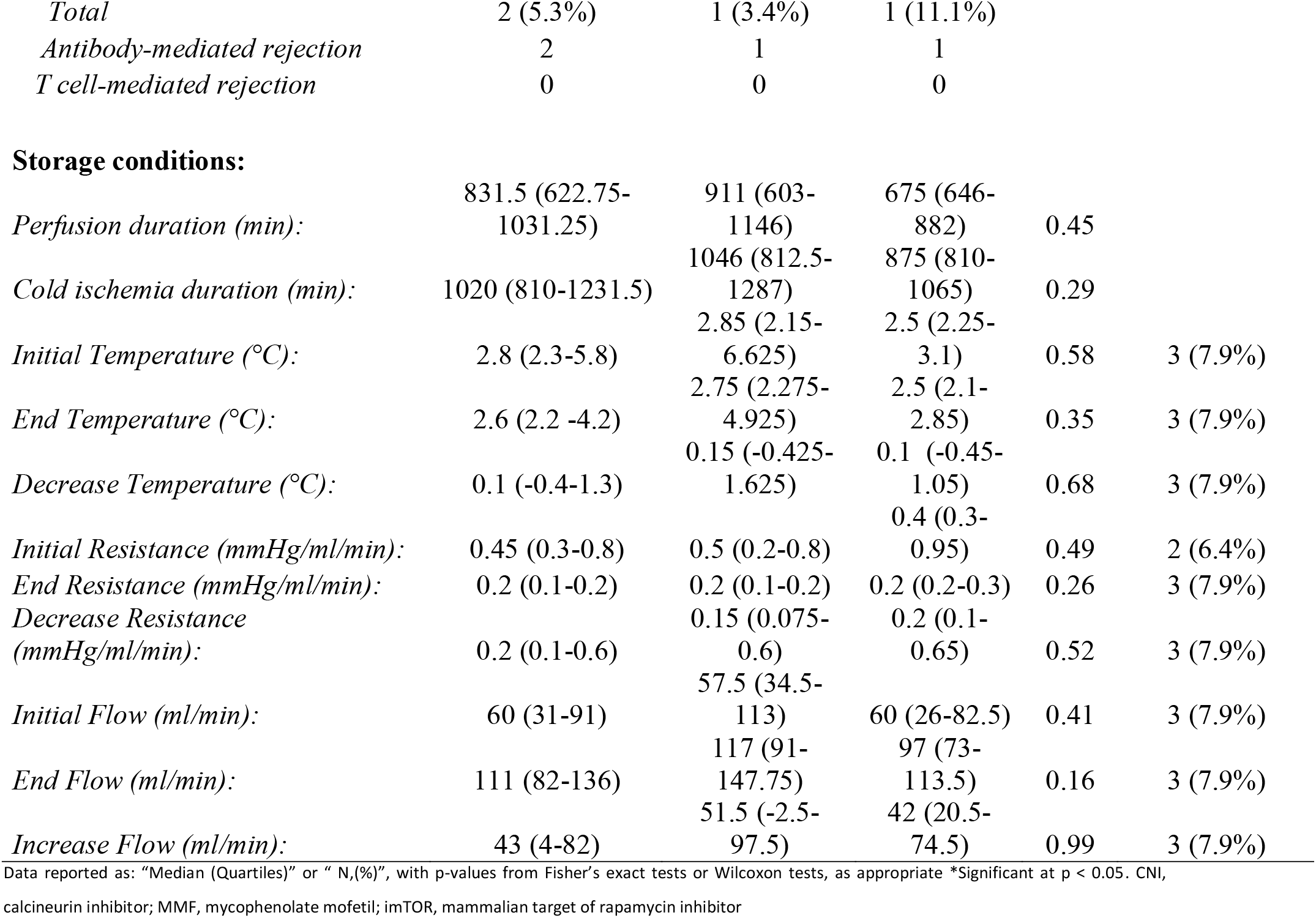
Characteristics of kidney graft donors, recipients and storage conditions

|  | Overall (n=38) | IGF (n=29) | non-IGF (n=9) | P | Unknown (N,(%)) |
| --- | --- | --- | --- | --- | --- |
| **Donor:** |  |  |  |  |  |
| Age (years): | 67.5 (61.75-75) | 65 (60.5-75) | 70 (67-77) | 0.28 |  |
| Sex (N,(%)): |  |  |  |  |  |
| F | 21 (55.3%) | 14 (48.3%) | 7 (77.8%) | 0.15 |  |
| M | 17 (44.8) | 15 (51.7 %) | 2 (22.2) |  |  |
| Donation after brain death (N,(%)): | 38 (100%) | 29 (100.0%) | 9 (100.0%) | 1 |  |
| Expanded Criteria donors (N,(%)): | 38 (100%) | 29 (100.0%) | 9 (100.0%) | 1 |  |
| **Recipient:** |  |  |  |  |  |
| Age (years): | 65.5 (57-72) | 65 (56.5-69) | 72 (61-73) | 0.20 |  |
| Sex (N,(%)): |  |  |  |  |  |
| F | 13 (34.2%) | 11 (37.9%) | 2 (22.2%) | 0.4562 |  |
| M | 25 (65.8 %) | 18 (62.1 %) | 7 (77.8 %) |  |  |
| Ethnicity (N,(%)): |  |  |  | 0.89 | 1 (2.6%) |
| Caucasian | 30 (78.9%) | 22 (75.9%) | 8 (88.9 %) |  |  |
| Afroamerican | 2 (5.3%) | 1 (3.4%) | 1 (11.1 %) |  |  |
| Other | 4 (10.5%) | 4 (13.8%) | 0 (0,0 %) |  |  |
| Renal disease : |  |  |  |  |  |
| Diabetes mellitus | 9 (23.7 %) | 6 (20.7%) | 3 (33.3 %) |  |  |
| Hypertension | 14 (36.8%) | 9 (31.0%) | 5 (55.6%) |  |  |
| Glomerulopathy | 2 (5.3%) | 1 (3.4%) | 1 (11.1 %) |  |  |
| Tubulointerstitial nephropathy | 3 (7.9%) | 1 (3.4%) | 2 (22.2%) |  |  |
| Polycystic kidney disease | 6 (15.8%) | 5 (17.2%) | 1 (11.1 %) |  |  |
| Other | 16 (42.1%) | 13 (44.8%) | 3 (33.3 %) |  |  |
| Number of Transplantation (N,(%)): |  |  |  | 1 |  |
| 1 | 35 (92.1%) | 26 (89.7%) | 9 (100.0%) |  |  |
| >1 | 3 (7.9%) | 3 (10.3%) | 0 (0.0%) |  |  |
| Dialysis before transplantation (N,(%)) : | 34 (89.5%) | 25 (86.2%) | 9 (100.0%) | 0.55 |  |
| Induction therapy (N,(%)): |  |  |  |  |  |
| Thymoglobulin | 11 (28.9%) | 10 (34.5%) | 1 (11.1%) |  |  |
| IL2R antibody | 27 (71.0%) | 19 (65.5%) | 8 (88.9%) |  |  |
| Maintenance therapy (N,(%)): |  |  |  |  |  |
| MMF-Corticosteroids-CNI | 32 (84.2%) | 24 (82.8%) | 8 (88.9%) |  |  |
| MMF-Corticosteroids | 3 (7.9%) | 2 (6.9%) | 1 (11.1%) |  |  |
| Corticosteroids-CNI-imTOR | 2 (5.3%) | 2 (6.9%) | 0 (0.0%) |  |  |
| Corticosteroids-CNI | 1 (2.6%) | 1 (3.4%) | 0 (0.0%) |  |  |
| Death (N,(%)): | 1 (2.6%) | 1 (3.4%) | 0 (0.0%) | 1 |  |
| Banff Classification (pre-implantation biopsies) (N,(%)): |  |  |  | 0.50 | 10 (26.3) |
| Normal | 17 (44.7%) | 12 (41.4%) | 5 (55.6%) |  |  |
| Acute tubular injury | 10 (26.3%) | 9 (31.0%) | 1 (11.1%) |  |  |
| Interstitial fibrosis/tubular atrophy | 1 (2.6%) | 1 (3.4%) | 0 (0.0%) |  |  |
| Graft rejection (N,(%)): |  |  |  |  |  |

|  |  |  |  |
| --- | --- | --- | --- |
| *Total* | 2 (5.3%) | 1 (3.4%) | 1 (11.1%) |
| *Antibody-mediated rejection* | 2 | 1 | 1 |
| *T cell-mediated rejection* | 0 | 0 | 0 |
| **Storage conditions:** |  |  |  |  |  |
| *Perfusion duration (min):* | 831.5 (622.75-1031.25) | 911 (603-1146) | 675 (646-882) | 0.45 |  |
| *Cold ischemia duration (min):* | 1020 (810-1231.5) | 1046 (812.5-1287) | 875 (810-1065) | 0.29 |  |
| *Initial Temperature (°C):* | 2.8 (2.3-5.8) | 2.85 (2.15-6.625) | 2.5 (2.25-3.1) | 0.58 | 3 (7.9%) |
| *End Temperature (°C):* | 2.6 (2.2 -4.2) | 2.75 (2.275-4.925) | 2.5 (2.1-2.85) | 0.35 | 3 (7.9%) |
| *Decrease Temperature (°C):* | 0.1 (-0.4-1.3) | 0.15 (-0.425-1.625) | 0.1 (-0.45-1.05) | 0.68 | 3 (7.9%) |
| *Initial Resistance (mmHg/ml/min):* | 0.45 (0.3-0.8) | 0.5 (0.2-0.8) | 0.4 (0.3-0.95) | 0.49 | 2 (6.4%) |
| *End Resistance (mmHg/ml/min):* | 0.2 (0.1-0.2) | 0.2 (0.1-0.2) | 0.2 (0.2-0.3) | 0.26 | 3 (7.9%) |
| *Decrease Resistance (mmHg/ml/min):* | 0.2 (0.1-0.6) | 0.15 (0.075-0.6) | 0.2 (0.1-0.65) | 0.52 | 3 (7.9%) |
| *Initial Flow (ml/min):* | 60 (31-91) | 57.5 (34.5-113) | 60 (26-82.5) | 0.41 | 3 (7.9%) |
| *End Flow (ml/min):* | 111 (82-136) | 117 (91-147.75) | 97 (73-113.5) | 0.16 | 3 (7.9%) |
| *Increase Flow (ml/min):* | 43 (4-82) | 51.5 (-2.5-97.5) | 42 (20.5-74.5) | 0.99 | 3 (7.9%) |

Data reported as: "Median (Quartiles)" or "N,(%)", with p-values from Fisher's exact tests or Wilcoxon tests, as appropriate \*Significant at p < 0.05. CNI, calcineurin inhibitor; MMF, mycophenolate mofetil; imTOR, mammalian target of rapamycin inhibitor

### 2. Perfusate metabolomic content and transcriptional expression of tubular transporters

#### 2.1. Metabolomic content of graft perfusion fluid

In the 35 perfusion liquids available, 72 different metabolites were identified, 66 with LC-MS/MS (Supplementary Table 2) and 6 with COBAS analyzer. All of them were present in each sample, with a few exceptions (Table 2). Twenty-nine of them were already present in the native KPS-1. At the end of perfusion, 6 were increased (inosine, guanosine, xanthosine, 5-methyl adenosine, tryptophan and riboflavin) and 10 were decreased (gluconic acid, methionine sulfoxide, glucosamine, oxidized glutathione, adenine, 2-Ketoisovaleric acid, 3-Methyl-2-oxovaleric acid, D-gluconic acid sodium salt, D-Ribose and deoxycytidine monophosphate) as compared to native KPS-1. Forty-three metabolites were exclusively found in the graft perfusates. These metabolites belong mainly to the amino-acids metabolism pathways (Supplementary Figure S1).

**Table 2:**
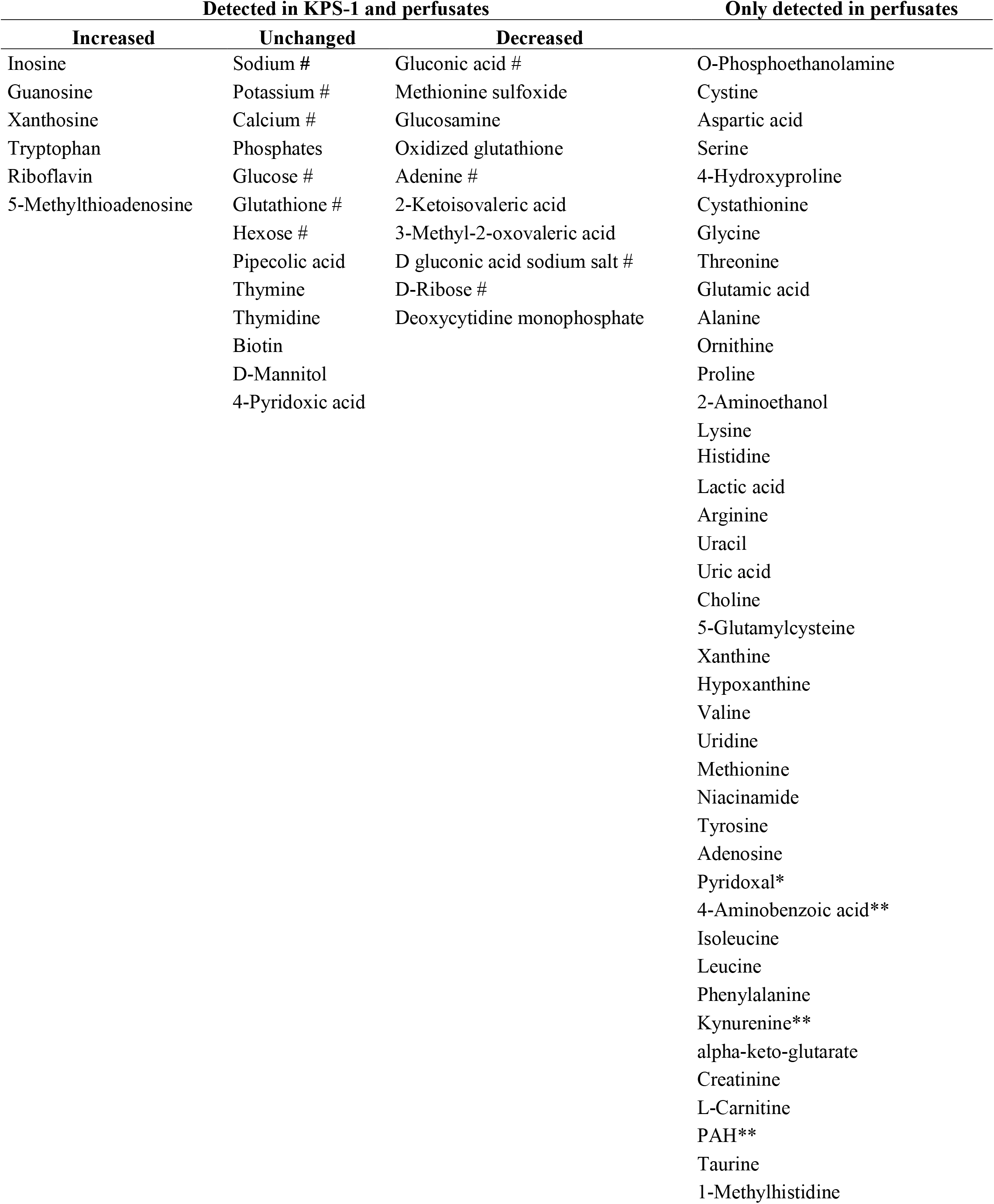
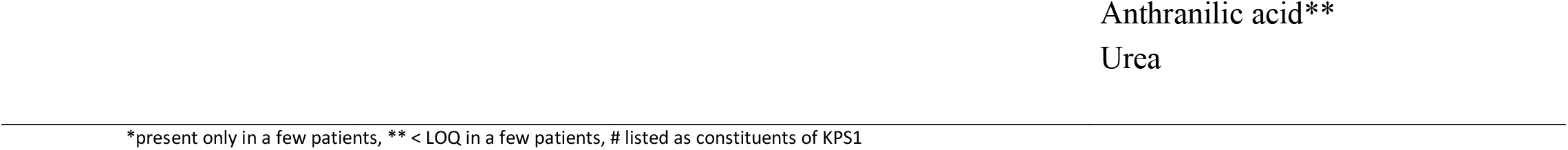
Metabolomic contents of commercial and end-of-perfusion KPS-1 preservation fluid

#### 2.2. Transcriptional expression of tubular transporters in pre-implantation biopsies

Thirty-four biopsies were available for RNA extraction. Eight were excluded: 2 because of RNA yield and 6 because they contained renal medulla. The mean RIN value for the 26 biopsies retained was 5.4 +/- 2.4. All the transporters of interest were identified in these biopsies. We found high expression correlations between them (Supplementary Figure S2).

### 3. Impact of perfusion duration on metabolomic profiles and transporter expression

#### 3.1. Metabolomics variations according to perfusion duration

Univariate analysis showed that 31 features were significantly (FDR<0.05) different between the perfusion duration groups (Figure 1). Twenty-three metabolites increased with longer perfusion durations (*e*.*g*. lactate or leucine) but 8 decreased (*e*.*g*. glutathione or inosine). PCA unsupervised analysis showed good separation of the scores between the two extreme groups (<12 h and >20 h), whereas group 2 overlapped with the others. Fifty-two percent of the variation was explained by the first and second components (Figure 2A). Similarly, the PLS-DA score plot (first two components) showed complete separation between groups 1 and 3, but overlap with group 2 (Figure 2B). Cross-validation showed good performances: goodness-of-fit (R^2^) = 76%, predictive cumulative variation (Q^2^) = 54% and accuracy = 41%. The model was significant according to the permutation test (p<0.05), supporting the absence of overfitting. The most important metabolites for the model are shown in Figure 2C. Significant pathways based on the important metabolites (VIP value > 0.8) are listed in Supplementary Figure S3.

**Figure 1:**
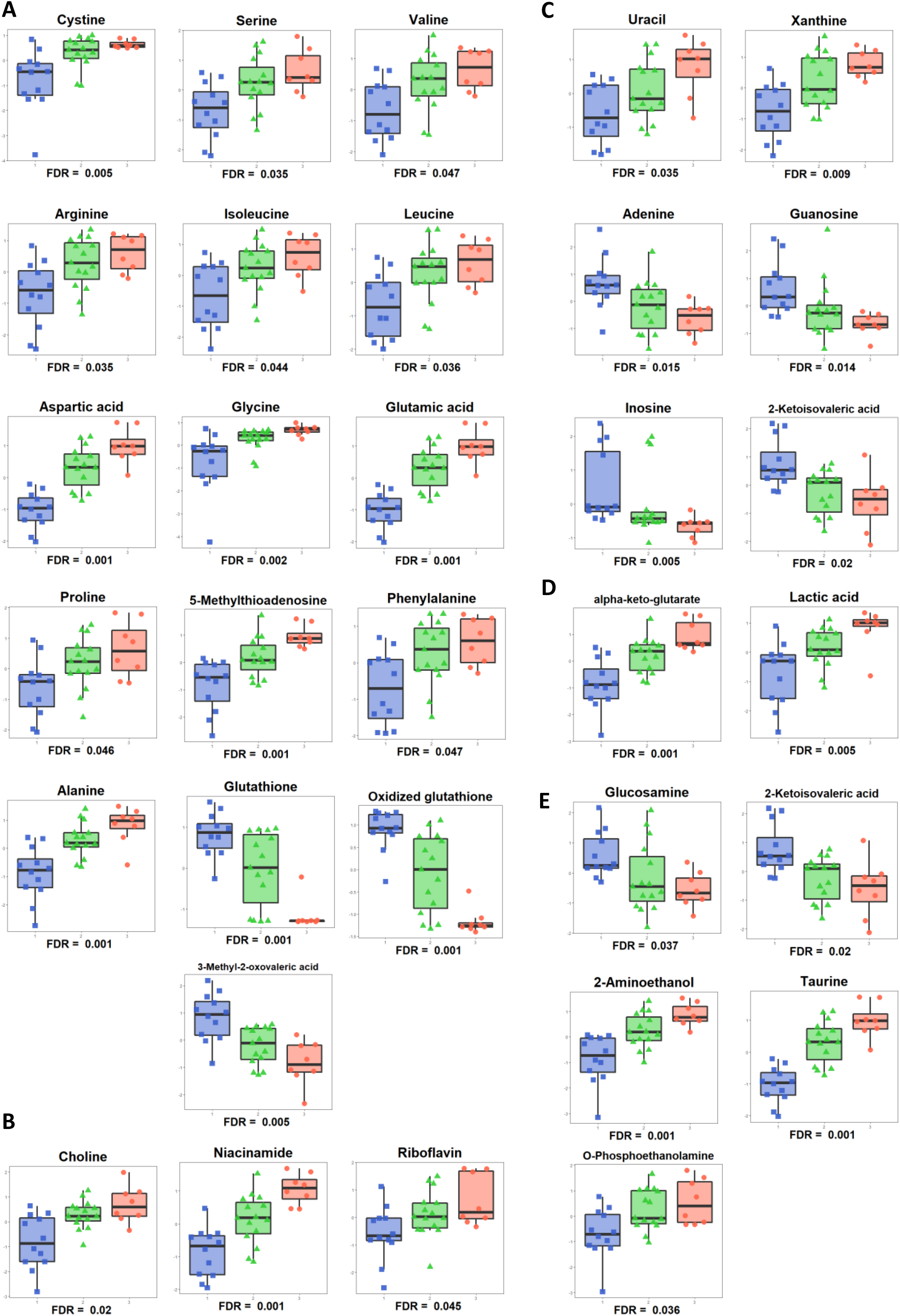
Univariate analysis of metabolites in perfusates according to perfusion duration. Only the metabolites with significant differences between groups are displayed in this figure. Comparisons were made using Kruskal-Wallis tests and adjusted for multiple testing by the FDR method. Group 1 (blue box): perfusion duration < 12 hours; Group 2 (green box): perfusion duration between 12 and 20 hours; Group 3 (red box): perfusion duration > 20 hours. Box plots are grouped according to compound classes: (A.) amino-acids and their metabolites, (B.) vitamins, (C.) nucleic acids and their metabolites, (D.) TCA Cycle and lactate, (E.) others.

**Figure 2:**
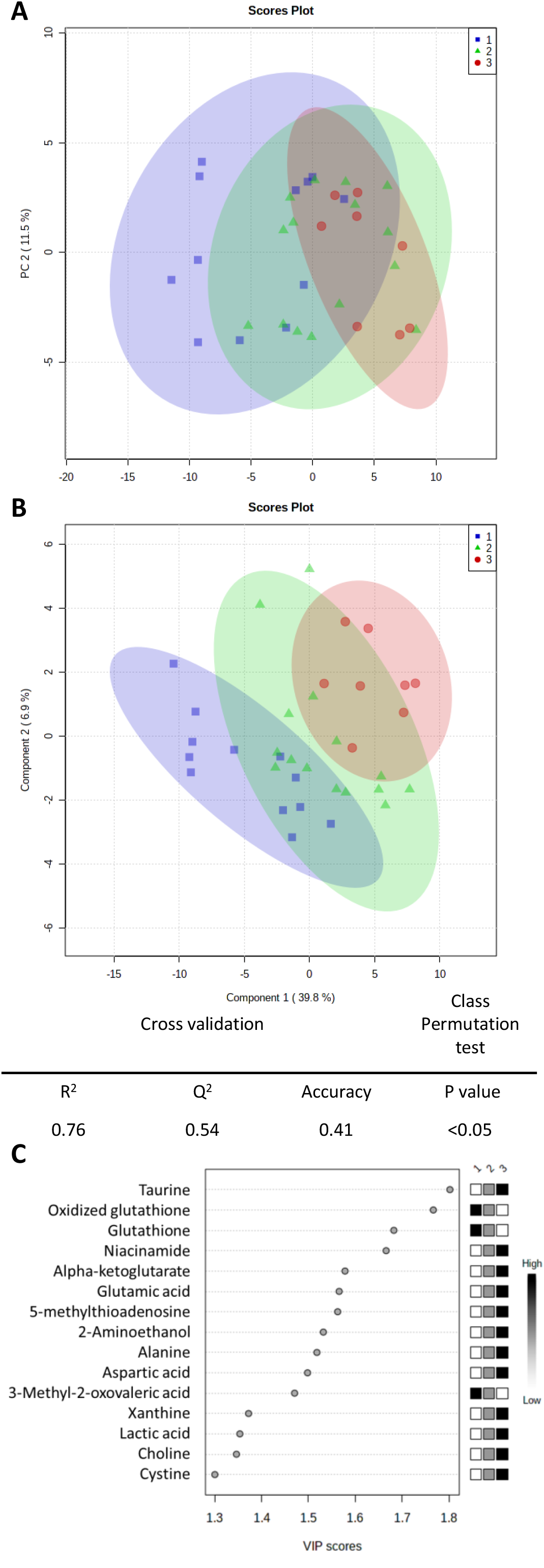
Multivariate analysis showing variations of metabolomic profiles according to perfusion duration. Group 1 (blue squares): perfusion duration < 12 hours; Group 2 (green triangles): perfusion duration between 12 and 20 hours; Group 3 (red dots): perfusion duration > 20 hours (A.) PCA scores plot showing separation between groups 1 and 3 but not with group 2. Principal component PC1 described 40% of the variation and PC2 11.5%. (B.) PLS-DA scores plot showing good separation between the 3 groups. (C.) Most important features for PLS-DA based on the (VIP) values; boxes on the right indicate the way of variation. PCA: Principal Component Analysis; PLS-DA: Partial Least Squares - Discriminant Analysis; VIP: Variable Influence on Projection.

#### 3.2. Transporter expression according to perfusion duration

The transcriptional expression of transporters according to the perfusion duration was not significantly different in univariate analysis. The PLS-DA model performance was: R^2^ = 34%, negative Q^2^ and accuracy = 30%, and the permutation test was not significant (Figure 3).

**Figure 3:**
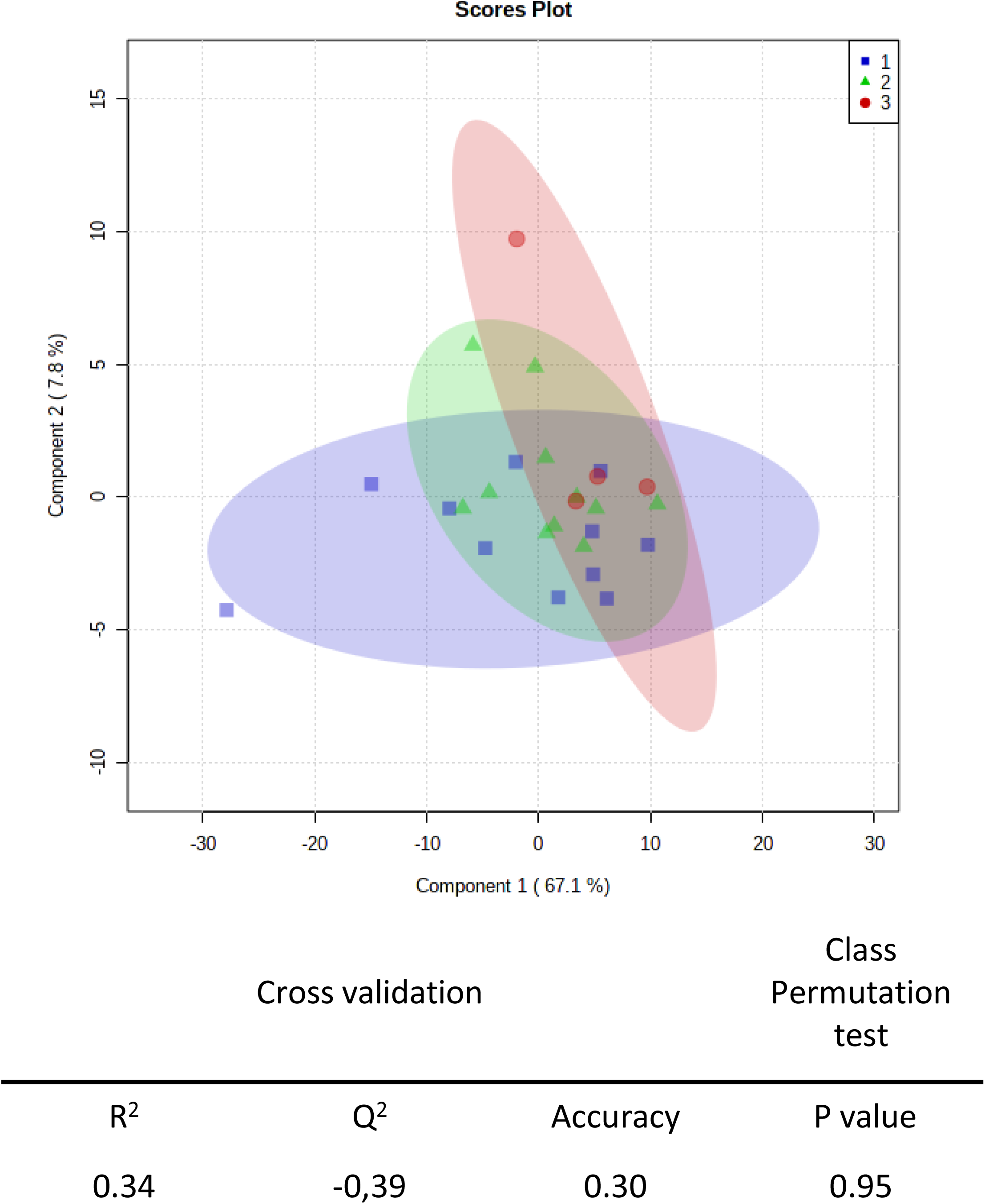
Variations of mRNA expression of tubule transporter according to perfusion duration. PLS-DA scores plot showed no separation between the 3 groups. PLS-DA model yielded poor performance with cross-validation (R^2^ = 34%, Q^2^ = -39% and accuracy = 30%) and the permutation test (p-value = 0.95). Group 1 (blue squares): perfusion duration < 12 hours; Group 2 (green triangles): perfusion duration between 12 and 20 hours; Group 3 (red dots): perfusion duration > 20 hours

### 4. Correlation between transporters expression and metabolomic content

Correlation between transporters expression and metabolomic content was investigated by PLS regression. The Clustered Image Maps (CIM) of the model is displayed in Figure 4. It does not show patterns of correlation between metabolites and transporters. The maximal positive and negative correlations were weak: 0.54 and -0.54, respectively.

**Figure 4:**
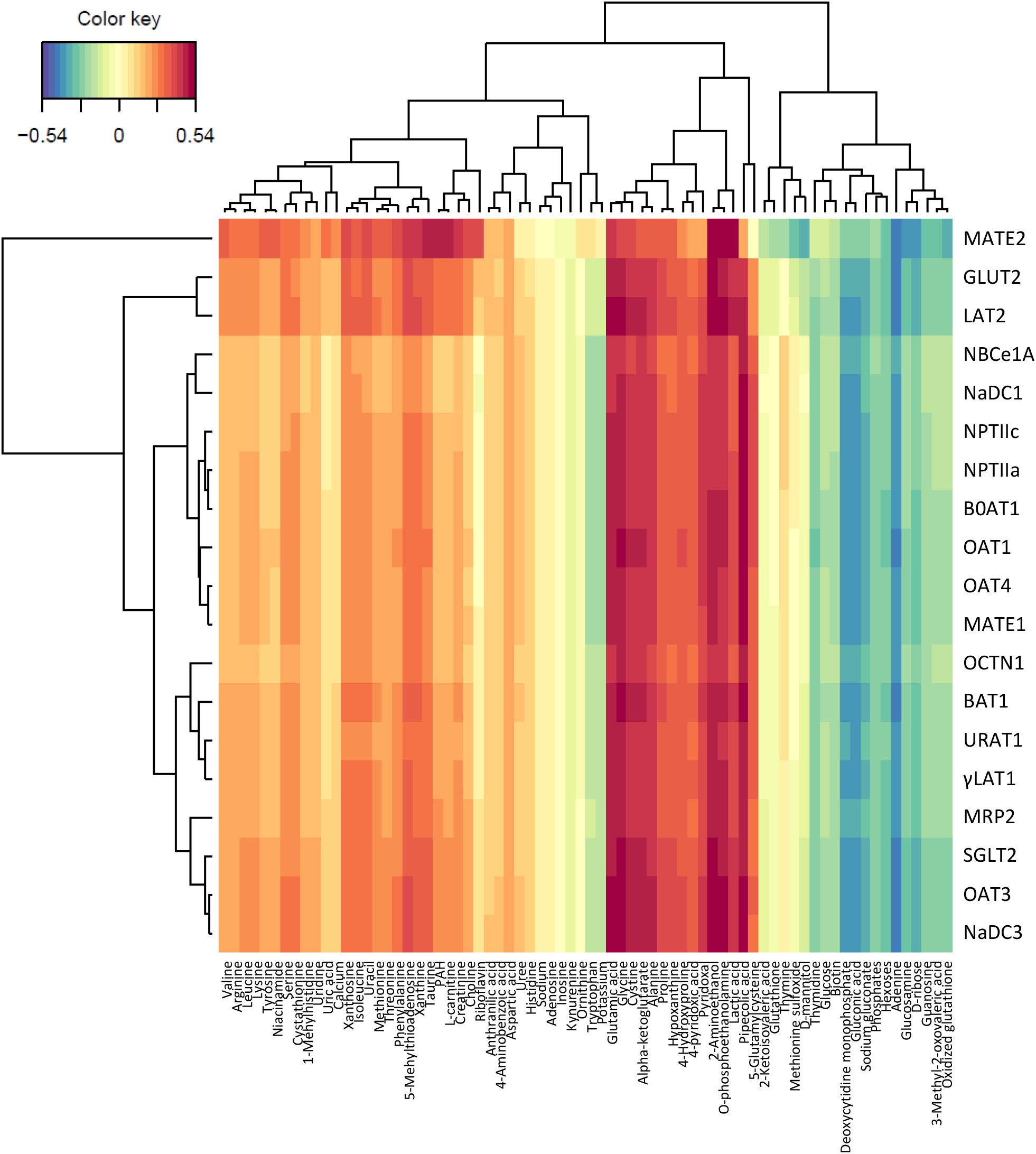
Clustered Image Maps of the relationships between transporters and metabolites (sPLS method). The red and blue colors indicate positive and negative correlations respectively, whereas yellow indicates no significant correlation. CIM: Clustered Image Maps; sPLS: sparse Partial Least Squares

### 5. Predictive biomarkers of graft outcome

Considering the low rate of patients with DGF (13.2%, n=5), we enhanced the statistical power of graft recovery analysis by comparing patients with and without IGF (serum creatinine > 250 µM on Day7, with or without dialysis). No significant differences were found between IGF (n=29) and non-IGF (n=9) recipients regarding the clinical parameters (*e*.*g*. age, sex, induction therapy), machine parameters recorded (temperature, resistive index, flow rate), cold ischemia time or biopsy pathological evaluation (Table 1).

#### 5.1. Metabolomics in assessing the quality of kidney grafts

Univariate analysis showed that 4-hydroxyproline (Fold Change (FC): 0.31, raw p-value: 0.13) and alanine (FC: 0.73, raw p-value: 0.15) tended to be lower in perfusion fluids of non-IGF patients (Figure 5A). PLS-DA allowed visual separation between IGF and non-IGF on the scoreplot of the first two components (Figure 5B), with correct accuracy (69 %) and R^2^ (36%), but negative Q^2^. The permutation test was not significant. Random forest analysis, a tree-based machine-learning algorithm adapted to small size datasets, was used to find a better model. The out-of bag error (samples wrongly predicted) was 23% for the overall dataset, but 100% of non-IGF samples were predicted as IGF, showing that the model was unable to predict graft recovery (Figure 5C).

**Figure 5:**
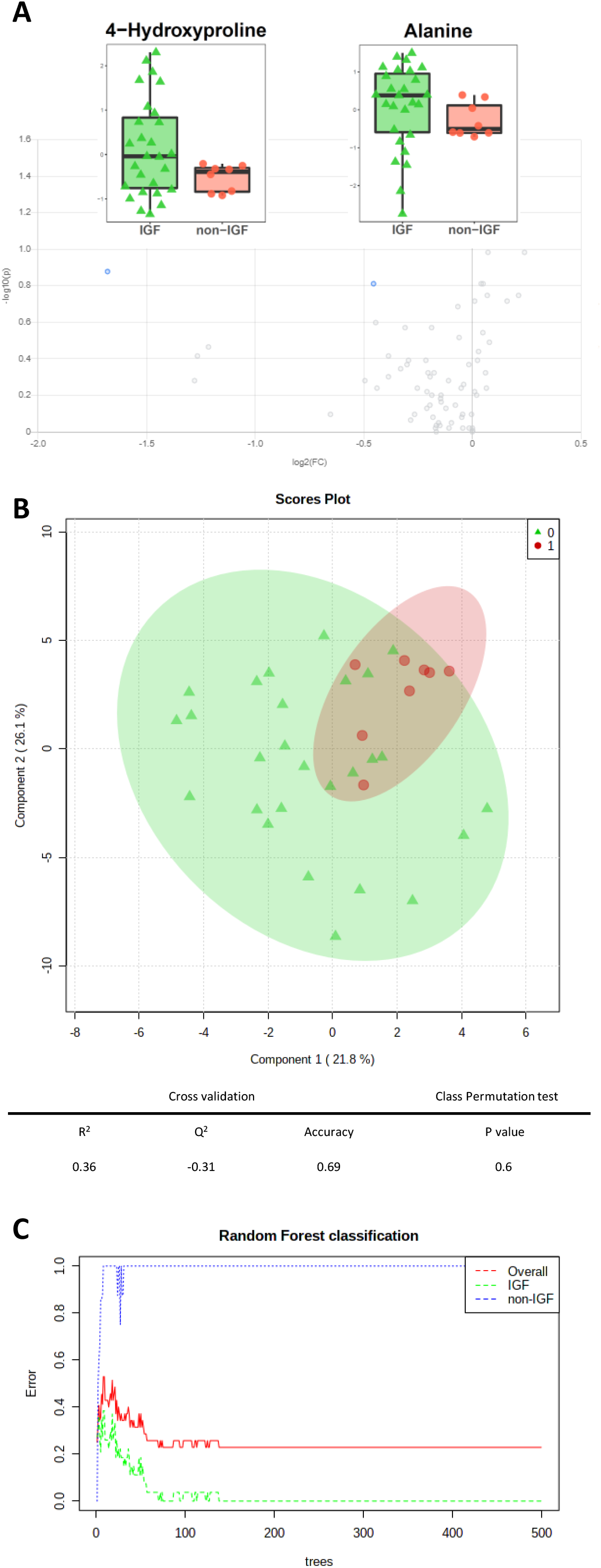
Assessment of IGF based on metabolomic profiles. Univariate analysis (A.) Volcano plot showing a decreased tendency of 4-Hydroxyproline and Alanine in non-IGF (red boxes) versus IGF (Immediate Graft Function; green boxes) patients (FC threshold of 1.2 and raw p-value of 0.2). Multivariate analysis (B.) The PLS-DA scores plot shows incomplete separation between groups with correct accuracy (70 %) and R^2^ (36%), but poor predictability (Q^2^ = -0.31). The permutation test was not significant (p = 0.6). (C.) Random Forest (RF) Classification showing the error rate for the overall dataset and for each class. PLS-DA; Partial Least Squares - Discriminant Analysis.

#### 5.2. Transcriptomic extraction of transporters according to graft quality

Univariate analysis did not identify any transporter able to discriminate between IGF and non-IGF (data not shown). Similarly, PLS-DA failed to predict graft recovery in our cohort, as evidenced by an overlap between the two groups (Figure 6A): the yielded R^2^=26% and accuracy = 69%, with a negative Q^2^. The permutation test was not significant. Random forest analysis showed poor prediction of non-IGF patients: 5 of the 6 non-IGF cases were misclassified as IGF (Figure 6B).

**Figure 6:**
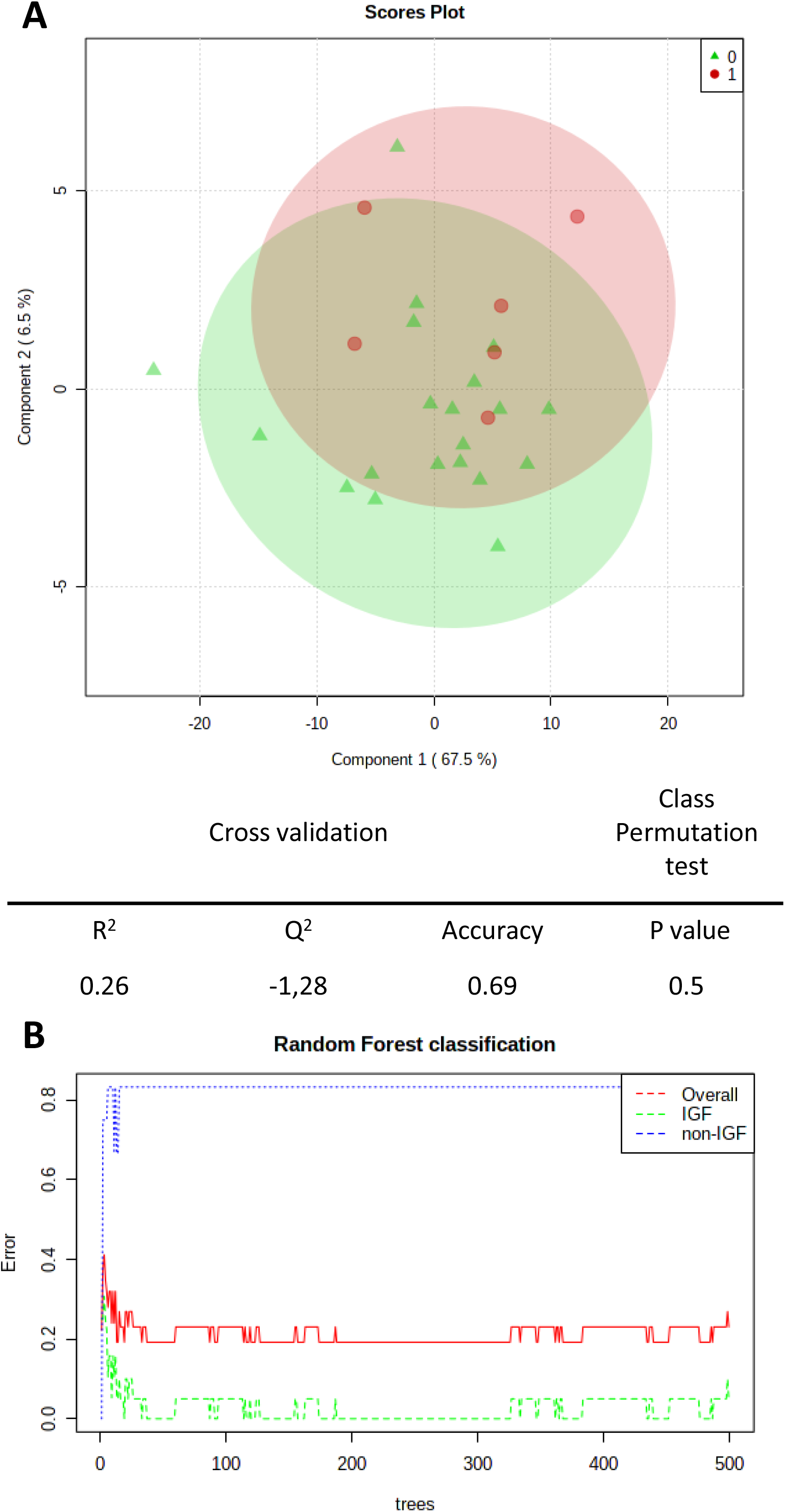
Assessment of IGF based on mRNA expression of tubule transporters. Multivariate analysis (A.) PLS-DA scores plot showing an overlap between graft recovery statuses and poor performance diagnostics (R^2^=26%, accuracy = 69% and Q^2^ = -1.28). The permutation test was not significant (p = 0.5) (B.) Random Forest (RF) Classification showing the overall error rate and for each class: the model has a poor predictability for non-IGF patients. PLS-DA; Partial Least Squares - Discriminant Analysis.

## Discussion

This clinical study aimed to better characterize the metabolic variations occurring during organ preservation on HMP through metabolic profiling of the fluid collected at the end of perfusion. We observed marked modifications of the metabolomic content, with regard to the native preservation fluid (KPS-1^®^), but also as a function of perfusion duration, which is a surrogate of the cold ischemia time duration. We also investigated the transcriptional expression of tubular transporters to explore if variations in their activity could be linked with metabolic variations seen during machine perfusion. However, we did not observe any particular pattern between the expression of any transporter and the metabolic profiles or perfusion duration. Finally, we evaluated the predictive ability of perfusate metabolites and tubular transporters mRNA expression as potential biomarkers of graft function by comparing IGF and non-IGF patients and found none.

### Maintained metabolic activity of grafts stored on perfusion machine

We determined the relative concentrations of 72 metabolites, some of which, to our knowledge, had never been studied in this context. We observed a marked difference in metabolomic profiles between the perfusate collected at the end of perfusion and the native fluid KPS-1 (Table 2). We found 29 metabolites in KPS-1, which is more than its theoretical composition. Among these, some showed decreased quantities at the end of perfusion, suggesting they were reabsorbed or consumed by the kidney, while others increased (Table 2). The 43 other metabolites, mainly amino-acids (Supplementary Figure 1), were exclusively detected in the fluid at the end of perfusion. Metabolites that appeared, or increased from basal value, were released by the graft, which can be related to sustained metabolic activity, ischemic damages, or both.

### Modifications occur during preservation on HMP

Understanding ischemic phenomena occurring during graft preservation remains a leading issue in renal transplantation for the improvement of graft management. However, our understanding of the metabolic activity occurring during kidney preservation on HMP is still partial. Only a few studies have analyzed kidney graft *in situ* or perfusate metabolomics^10–12,22^. They aimed to identify early predictive biomarkers of graft function and investigate the underlying mechanisms. Our work attempted to consolidate and complement previous studies by: (i) the use of LC-MS/MS instead of NMR^10–12^; (ii) the exploration of human kidneys instead of pig^10^ or rabbit models^22^; and (iii) the exploration of longer perfusion times^11,12^. Among the 72 metabolites analyzed, the concentration of 8 them significantly decreased when the perfusion time increased (Figure 1), including glutathione and oxidized glutathione. This decrease had already been described by ^1^H-NMR analyses of perfusates in both humans and pig^10–12^. Glutathione is involved in free radicals scavenging and its decrease sensitizes the graft to IRI. We also found an increased level of lactic acid, choline and some amino-acids (*e*.*g*. valine, alanine, glycine and glutamic acid) as a result of longer perfusion. These observations are consistent with previous results obtained by Bon *et al*., who reported an increase in concentrations of these metabolites in porcine kidney perfusates between 2h and 22h perfusion^10^. Increased lactic acid simply reflects anaerobic glycolysis occurring in a hypoxic environment and increased amino-acids may reflect intracellular release. Most of our findings are consistent with those previously published, except for some. For example, the level of glucose and inosine increased with perfusion time in the study by Guy *et al*., whereas it was unchanged and decreased in ours. However, these authors explored two early time points: 45 min vs. 4 hours, while the first quartile of our perfusion duration was already about 10 hours. The usefulness of following the kinetics of metabolites has been demonstrated recently, using solid phase microextraction and LC/MS for *in situ* kidney metabolomics analysis at five sequential time points in a rabbit model: immediately following removal of the donor’s kidneys and after 2, 4, 6 and 21 hours of SCS^22^. The authors found that metabolites related to various metabolic pathways, including amino-acids and purine metabolism, significantly increased during the first hours of kidney preservation, whereas a decrease occurred with longer perfusion durations. However, their methodology (*in situ*, limited number of samples, SCS conservation) hampers the direct comparison with our results. Nevertheless, we also observed at the extra-tissue level that the amounts of some intermediates of purine metabolism such as adenine, inosine and guanosine decreased, while that of xanthine increased, with perfusion time. The results observed here most likely illustrate that kidneys with longer ischemia consume more adenine and inosine and produce more xanthine in response to ATP deprivation. To the best of our knowledge, other metabolites (*e*.*g*. taurine, niacinamide or glucosamine) and their association with perfusion duration had never been studied in this context.

### Weak correlation between metabolites in the perfusate and tubular transporters

The metabolomic signatures observed over perfusion time reflect sustained metabolic activities by kidneys stored on HMP, but the underlying mechanisms are currently poorly understood. Some metabolites are physiologically reabsorbed or secreted through specific membrane tubular transporters^15^. Their sensitivities to ischemia^15,23^ and potentially altered activity could explain a part of the *ex situ* metabolomic variations observed. A decrease at the mRNA and protein levels of *SLC22A6* (OAT1) and *SLC22A8* (OAT3), involved in the uptake of organic anions, was observed after 30 minutes of ischemia in the rat^13^ and similar results were found for *SLC22A1* (OCT1) and *SLC22A2* (OCT2), involved in organic cation uptake, also in a rat model^14^. Considering the limited quantity and quality of RNA we chose a TLDA approach for transcriptional expression determination, since it only requires small quantities of RNA and is able to amplify small cDNA fragments. We successfully determined the transcriptional expression of all targeted genes. However, no association was observed between transporter expression and perfusion duration (Figure 3). Previous works suggest that the downregulation of membrane transporters occurs since only after 30 min of warm ischemia^23^. In our study, some transporter alterations may have occurred before the biopsy, which was done after the warm ischemia period and after at least 176 min of perfusion. We might have confirmed this hypothesis with non-ischemic biopsy controls, but we could not obtain any. It is also possible that some transporters are not affected by ischemia during preservation by HMP.

We did not find any particular correlation with the metabolomic profile in the perfusion fluid, even for well-known transporter/substrate couples (*e*.*g*., SGLT2/glucose or OAT1/PAH) and amino-acid transport systems (Figure 4). In HMP, the perfusion fluid irrigates both the basolateral and apical poles of the tubule, whereas the function of transporters and thus the identification of variations reflecting their activity is highly dependent on a polarized environment. Moreover, the transcriptional expression of transporters alone cannot perfectly reflect their real activity, but we could not evaluate their protein expression (because of too low amounts of proteins) or cellular localization (due to histological fixation). Globally, these results call for further studies, evaluating both the intra- and extra-tissue metabolome in relation to the expression, localization and functions of transporters (e.g. based on transporter-specific labeled substrates) as a function of ischemia time, to uncover potential relationships between transporters and metabolites during kidney graft perfusion. However, it is worth mentioning that such studies may still be confounded by injury-induced cellular release of metabolites.

### Metabolomic signatures and expression of tubular transporters do not predict IGF

The present study also aimed to identify non-invasive biomarkers predictive of graft outcomes that could be measured in the pre-implantation period. Actually, reliable biomarkers would be instrumental to optimize patient management and graft outcome. Assessment of graft quality in the transplantation period is currently based on perfusion parameters such as the restrictive index or the flow rate, and pathological evaluation of a preimplantation biopsy. However, these indicators are not sufficiently reliable to safely discard a graft. Accordingly, in our study, no indicators predicted post-transplant graft function (data not shown). Rapid determination of the perfusate metabolomic profile in the preimplantation period would be convenient and adapted to clinical routine. The proof of concept was brought by Bon *et al*. in a porcine renal auto-transplantation model where valine, alanine, glutathione and glutamate concentrations in the perfusion fluid were correlated with serum creatinine at 3 months^10^. Guy *et al*. also found in a human cohort that glucose and inosine concentrations were lower in the perfusate of DGF kidneys, while leucine was higher^11^. In our study, we did not identify any metabolite or multivariate model predictive of graft recovery (Figure 5). We did not replicate previous results, maybe due to the differences of perfusion durations explored and given that in the study by Guy *et al*., cadaveric kidneys arrived at their unit on SCS before being transferred to HMP, whereas in our study the kidneys retrieved from each donor were put directly on HMP. Moreover, the metabolites identified previously were not the same between the two studies.

The relative heterogeneity and the small size of our cohort probably limited the predictive ability of the metabolome. Also, the low DGF rate (13.2%) of ECD donors’ kidneys preserved with the LifePort^®^ Kidney Transporter in our cohort (as compared with a DGF rate of 30 % for SCS in France in 2017^24^) supported the utility of HMP for such donors, but limited the statistical power of our study. However, our negative result is in line with a recent systematic review that highlighted the lack of accuracy and hindsight of metabolomics in human kidney graft perfusates^16^. The evaluation of perfusate metabolomics variations might still be relevant^25^, since it recently suggested a higher *de novo* metabolic activity of kidneys preserved on machine versus SCS^26^, or it could highlight the metabolic variations occurring with oxygen supplementation^27,28^ or pharmacological agents^29,30^.

Finally, we aimed to determine if the expression of tubular transporters in our cohort could predict early post-transplant graft outcomes. To the best of our knowledge, no study has yet been conducted in this regard. Our results do not support evaluating transporters transcriptional expression at the end of kidney graft HMP, with the aim of predicting graft function.

## Conclusion

In summary, we did not find any predictive biomarkers of graft function in the perfusate metabolome or among tubular transporter mRNAs of human kidneys stored on HMP. However, we observed marked differences between the metabolomics profiles collected at the end of perfusion and the perfusion liquid initial composition, which reflects persistence of a metabolic activity during HMP. Moreover, the concentration of many metabolites was modified after the longer perfusions, mostly in agreement with the literature but also for some metabolites that have never been studied in this context so far. The transcriptional expression of 40 membrane transporters determined at the same time was not correlated with the variations of these metabolites or with perfusion time. We suggest conducting further translational studies, to evaluate the ratio of tissue-to-perfusion fluid concentrations of metabolites as well as tubular transporter activity to decipher the deleterious mechanisms associated with ischemia in the pre-implantation period.

## Supporting information

Supplemental Digital Content

## Data Availability

Data are available upon request

## Acknowledgments

The authors thank the Nouvelle Aquitaine Region, INSERM and the Organ Recovery System for their financial support. The authors are grateful to Karen Poole for correcting English and Etienne Broggi for his valuable input during the initiation of the project.

## ABBREVIATIONS PAGE

ABC: ATP-binding cassette
CIM: clustered image maps
DGF: delayed graft function
ECD: expanded criteria donors
FC: fold change
FDR: false discovery rate
HMP: hypothermic machine perfusion
IGF: immediate graft function
IRI: ischemia-reperfusion injury
LC-MS/MS: liquid chromatography/tandem mass spectrometry
NMR: nuclear magnetic resonance
PCA: principal component analysis
PLS: partial least squares
PLS-DA: partial least squares discriminant analysis
RF: random forest
RIN: RNA integrity number
SCS: static cold storage
SLC: solute carrier

## Bibliography

1. World Health Organization. Projections of mortality and causes of death, 2016 to 2060. Accessed May 13, 2021. https://www.who.int/healthinfo/global_burden_disease/projections/en/

2. Liyanage T, Ninomiya T, Jha V, et al. Worldwide access to treatment for end-stage kidney disease: a systematic review. The Lancet. 2015;385(9981):1975–1982. doi:10.1016/S0140-6736(14)61601-9

3. Wong G, Teixeira-Pinto A, Chapman JR, et al. The Impact of Total Ischemic Time, Donor Age and the Pathway of Donor Death on Graft Outcomes After Deceased Donor Kidney Transplantation. Transplantation. 2017;101(6):1152–1158. doi:10.1097/TP.0000000000001351

4. Chen C-C, Chapman WC, Hanto DW. Ischemia-reperfusion injury in kidney transplantation. Front Biosci (Elite Ed). 2015;7:117–134.

5. Gill J, Dong J, Rose C, Gill JS. The risk of allograft failure and the survival benefit of kidney transplantation are complicated by delayed graft function. Kidney Int. 2016;89(6):1331–1336. doi:10.1016/j.kint.2016.01.028

6. Nieuwenhuijs-Moeke GJ, Pischke SE, Berger SP, et al. Ischemia and Reperfusion Injury in Kidney Transplantation: Relevant Mechanisms in Injury and Repair. Journal of Clinical Medicine. 2020;9(1):253. doi:10.3390/jcm9010253

7. Salvadori M, Rosso G, Bertoni E. Update on ischemia-reperfusion injury in kidney transplantation: Pathogenesis and treatment. World J Transplant. 2015;5(2):52–67. doi:10.5500/wjt.v5.i2.52

8. Peng P, Ding Z, He Y, Zhang J, Wang X, Yang Z. Hypothermic Machine Perfusion Versus Static Cold Storage in Deceased Donor Kidney Transplantation: A Systematic Review and Meta-Analysis of Randomized Controlled Trials. Artif Organs. 2019;43(5):478–489. doi:10.1111/aor.13364

9. Barin-Le Guellec C, Largeau B, Bon D, Marquet P, Hauet T. Ischemia/reperfusion-associated tubular cells injury in renal transplantation: Can metabolomics inform about mechanisms and help identify new therapeutic targets? Pharmacol Res. 2018;129:34–43. doi:10.1016/j.phrs.2017.12.032

10. Bon D, Billault C, Claire B, et al. Analysis of perfusates during hypothermic machine perfusion by NMR spectroscopy: a potential tool for predicting kidney graft outcome. Transplantation. 2014;97(8):810–816. doi:10.1097/TP.0000000000000046

11. Guy AJ, Nath J, Cobbold M, et al. Metabolomic analysis of perfusate during hypothermic machine perfusion of human cadaveric kidneys. Transplantation. 2015;99(4):754–759. doi:10.1097/TP.0000000000000398

12. Nath J, Guy A, Smith TB, et al. Metabolomic Perfusate Analysis during Kidney Machine Perfusion: The Pig Provides an Appropriate Model for Human Studies. PLoS One. 2014;9(12). doi:10.1371/journal.pone.0114818

13. Matsuzaki T, Watanabe H, Yoshitome K, et al. Downregulation of organic anion transporters in rat kidney under ischemia/reperfusion-induced acute [corrected] renal failure. Kidney Int. 2007;71(6):539–547. doi:10.1038/sj.ki.5002104

14. Matsuzaki T, Morisaki T, Sugimoto W, et al. Altered pharmacokinetics of cationic drugs caused by down-regulation of renal rat organic cation transporter 2 (Slc22a2) and rat multidrug and toxin extrusion 1 (Slc47a1) in ischemia/reperfusion-induced acute kidney injury. Drug Metab Dispos. 2008;36(4):649–654. doi:10.1124/dmd.107.019869

15. Faucher Q, Alarcan H, Marquet P, Barin-Le Guellec C. Effects of Ischemia-Reperfusion on Tubular Cell Membrane Transporters and Consequences in Kidney Transplantation. J Clin Med. 2020;9(8). doi:10.3390/jcm9082610

16. Guzzi F, Knight SR, Ploeg RJ, Hunter JP. A systematic review to identify whether perfusate biomarkers produced during hypothermic machine perfusion can predict graft outcomes in kidney transplantation. Transplant International. 2020;33(6):590–602. doi:https://doi.org/10.1111/tri.13593

17. Wang Z, Lyu Z, Pan L, Zeng G, Randhawa P. Defining housekeeping genes suitable for RNA-seq analysis of the human allograft kidney biopsy tissue. BMC Med Genomics. 2019;12(1):86. doi:10.1186/s12920-019-0538-z

18. Schmittgen TD, Livak KJ. Analyzing real-time PCR data by the comparative C(T) method. Nat Protoc. 2008;3(6):1101–1108. doi:10.1038/nprot.2008.73

19. Vandesompele J, De Preter K, Pattyn F, et al. Accurate normalization of real-time quantitative RT-PCR data by geometric averaging of multiple internal control genes. Genome Biol. 2002;3(7):RESEARCH0034. doi:10.1186/gb-2002-3-7-research0034

20. Andersen CL, Jensen JL, Ørntoft TF. Normalization of real-time quantitative reverse transcription-PCR data: a model-based variance estimation approach to identify genes suited for normalization, applied to bladder and colon cancer data sets. Cancer Res. 2004;64(15):5245–5250. doi:10.1158/0008-5472.CAN-04-0496

21. Chong J, Wishart DS, Xia J. Using MetaboAnalyst 4.0 for Comprehensive and Integrative Metabolomics Data Analysis. Curr Protoc Bioinformatics. 2019;68(1):e86. doi:10.1002/cpbi.86

22. Stryjak I, Warmuzinska N, Bogusiewicz J, Luczykowski K, Bojko B. Monitoring of the influence of long-term oxidative stress and ischemia on the condition of kidney using solid phase microextraction chemical biopsy coupled with liquid chromatography high resolution mass spectrometry. J Sep Sci. Published online February 18, 2020. doi:10.1002/jssc.202000032

23. Kwon O, Wang W-W, Miller S. Renal organic anion transporter 1 is maldistributed and diminishes in proximal tubule cells but increases in vasculature after ischemia and reperfusion. Am J Physiol Renal Physiol. 2008;295(6):F1807–1816. doi:10.1152/ajprenal.90409.2008

24. Agence de la biomédecine - Le rapport annuel médical et scientifique 2017. Accessed October 15, 2019. https://www.agence-biomedecine.fr/annexes/bilan2017/accueil.htm

25. Kvietkauskas M, Zitkute V, Leber B, Strupas K, Stiegler P, Schemmer P. The Role of Metabolomics in Current Concepts of Organ Preservation. International Journal of Molecular Sciences. 2020;21(18):6607. doi:10.3390/ijms21186607

26. Nath J, Smith TB, Patel K, et al. Metabolic differences between cold stored and machine perfused porcine kidneys: A 1H NMR based study. Cryobiology. 2017;74:115–120. doi:10.1016/j.cryobiol.2016.11.006

27. Darius T, Vergauwen M, Smith TB, et al. Influence of Different Partial Pressures of Oxygen During Continuous Hypothermic Machine Perfusion in a Pig Kidney Ischemia-reperfusion Autotransplant Model. Transplantation. 2020;104(4):731–743. doi:10.1097/TP.0000000000003051

28. Patel K, Smith TB, Neil DAH, et al. The Effects of Oxygenation on Ex Vivo Kidneys Undergoing Hypothermic Machine Perfusion. Transplantation. 2019;103(2):314–322. doi:10.1097/TP.0000000000002542

29. Hosgood SA, Hoff M, Nicholson ML. Treatment of transplant kidneys during machine perfusion. Transpl Int. 2021;34(2):224–232. doi:10.1111/tri.13751

30. Franzin R, Stasi A, Fiorentino M, et al. Renal Delivery of Pharmacologic Agents During Machine Perfusion to Prevent Ischaemia-Reperfusion Injury: From Murine Model to Clinical Trials. Front Immunol. 2021;12. doi:10.3389/fimmu.2021.673562

