## Supplemental Digital Content for "Perfusate metabolomics content and tubular transporters expression during kidney graft preservation by hypothermic machine perfusion"

**SDC, Materials and Methods:**

1. **Study design :**

This two year prospective study relied on 38 kidney transplants from the Orléans Regional Hospital and University Hospital of Tours (France), within the framework of the clinical research project named RENALIFE, registered under ClinicalTrials.gov (NCT03024229). Patients were included if they met the following selection criteria: Donor aged at least 18 years, expressed non-opposition, dead brain deceased, and met the expanded criteria for renal retrieval (a donor aged either more than 50 or 60 years old, with two of the following criteria: a history of high blood pressure, a plasmatic creatinine greater than or equal to 1.5 mg/L, death resulting from a stroke). Immediately after the kidneys had been retrieved from the donor, they were stored on HMP, LifePort® Kidney Transporter 1.0 (Organ Recovery Systems). The organ preservation solution used in this device was KPS-1® solution (Organ Recovery System). The parameters of perfusion machine (i.e. temperature, flow, resistance) were recorded during kidney conservation. When the receiver was ready for transplantation, the graft was removed from the machine. An aliquot of the perfusion fluid was taken and immediately centrifuged at 3000g for 10 min then stored at -20°C. Just before transplantation, a systematic graft biopsy was also performed with a 16 Gauge gun (directed in order to sample the renal cortex). The biopsy was split transversely into two fragments: the first one was fixed in Bouin's solution (picric acid, acetic acid, formalin) or in AFA (acetic alcohol formalin) depending on the date of inclusion and then included in paraffin to carry out pathological classification, according to the revised Banff classification 2013; the second fragment was stored at -20°C. Each recipient benefited from a clinical and biological follow-up, including daily creatininemia measurement until day 7 and a collection of clinical events, which was carried out during the entire follow-up period, i.e. 3 months post-transplantation. The primary endpoints for graft recovery were Immediate Graft Function (IGF), characterized by a creatininemia at day 7 ≤ 250 µmol/L without the necessity for dialysis and Delayed Graft Function (DGF), characterized by the necessity for dialysis within 7 days.

### 2. Reagents

Discovery HS F5-3 column (150 x 2.1 mm d.i., 3 µm) (#567503-U), 2-Isopropylmalic acid (#333115), α-ketoglutaric acid (#75890), D-gluconic acid sodium salt (#G9005), D-Mannitol (#M4125), D-Ribose (#R7500), L-Carnitine (#8.40092), Magnesium D-gluconate hydrate (#G9130), P-Aminohippuric acid (#A1422) and Taurine (#T0625) were purchased from Sigma-Aldrich. Creatinine (27910) was purchased from Fluka. Taqman Low Density Array® (TLDA) cards were purchased from Thermofisher. The list of probe sets and manufacturer's code for TaqMan probes are listed in Supplementary Table 1.

### 3. Metabolomics analysis of graft preservation samples

#### 3.1. LC-MS/MS-metabolomic analysis

Mass spectrometry analysis of perfusion fluids was performed using LCMS-8060 (Shimadzu) tandem mass spectrometer and the LC-MS/MS "Method Package for Cell Culture Profiling Ver.2" (Shimadzu). In order to analyze compounds not initially included in this kit, an infusion of pure substances was performed and the corresponding transitions were added to the list of compounds to analyze. Briefly, 200 µL of acetonitrile and 20 µL of internal standard (2-Isopropylmalic acid [0.5 mmol/L]) were added to 100 µL of preservative solution from each patient. After centrifugation at room temperature for 15 minutes at 15,000 g, the supernatant was diluted 1/10th in ultrapure water and transferred to a vial before injection of 3µL into the analytical system. For each transition analyzed, only well-defined chromatographic peaks were considered. Each sample was analyzed in duplicate. Quality controls were prepared by mixing an equal volume of each patient's sample and were injected three times in all the series. Metabolites with a coefficient variation (CV) > 30% in quality controls were not retained for subsequent metabolomics analysis. Native KPS-1 was also injected to determine its basal metabolic composition. Raw spectrometric data sets were normalized in relation to the intensity of 2-Isopropylmalic acid as internal standard. Regarding the LC-MS/MS parameters: Chromatographic separation of the compounds was carried out with a Discovery HS F5-3 column (150 x 2.1 mm d.i., 3 µm) column thermostated at 40°C, at a flow rate of 350 µL/min according to the following linear mobile phase gradient: 0-1.4 min : 0% B ; 1.4-3.5 min : 0 to 25% B ; 3.5-7.5 min ; 25 to 35% B ; 7.5 to 10.3 min : 35 to 95% B; 10.3-13.7: 95%B; 13.7-13.8 min: 95 to 0% B; 13.8-17 min: 0%B where A is a 0.1% solution of formic acid in water and B is a 0.1% solution of formic acid in acetonitrile. Detection was performed by electrospray ionization using the following source conditions: Nebulizing gas flow rate; 3 mL/min; Drying gas flow rate: 10 mL/min; Heating gas flow rate: 10 mL/min; Interface temperature: 300°C; Desolvation line temperature: 250°C; block heater temperature: 400°C; Compounds were detected in scheduled MRM (Multi Reaction Monitoring) mode alternately in positive and negative modes, used for compound identification: one transition for quantification and a minimum of one transition for confirmation and relative retention time (Supplementary Table 2).

#### 3.2. **Assessment of other biochemical compounds**

COBAS 6000 analyzer (Roche Diagnostics) was used to determine sodium, potassium, calcium, phosphate, chloride, bicarbonate, urea, creatinine and glucose concentrations. The quantifiable features were then added to the list of metabolites measured by LC-MS/MS for subsequent analyses.

### 4. Transcriptional expression of tubular transporter

Frozen pre-implantation biopsies were used for RNA extraction, using the Nucleospin RNA/Protein kit (Macherey-Nagel), according to the manufacturer's instructions. RNA was then quantified using Qubit 4.0 with the Qubit RNA HS Assay kit (Thermofisher). RNA quality was assessed by RNA Integrity Number (RIN), determined with a Bioanalyzer 21 000 (Agilent). Agilent RNA 6000 Pico or Agilent RNA 6000 Nano kits were used according to RNA concentration. 200ng of RNA were then reverse transcribed into complementary DNA (cDNA) using the High capacity cDNA Reverse Transcription kit (ThermoFisher), according to the manufacturer's instructions. The TLDA card used enabled the quantification of 35 membrane tubular transporters , 3 aquaporins, 2 Na/K-ATPase subunits, and 4 housekeeping genes candidates (*NME4*, *CHFR*, *C16ORF62* and *NASP)* chosen according to the literature. For each sample, a mix containing all the cDNA diluted in RNAse-free water q.s. 55µL and 55 µL TaqMan® Universal Master Mix II was prepared. 100 µL of this reaction mix was then loaded onto each slot of the TLDA card. After double centrifugation at 1,200 rpm for 1 minute, the card was sealed and subsequently analyzed using Polymerase Chain Reaction (PCR) QuantStudio 12K with mix-UNG Amperase following the program conditions: 50°C for 2 min, 95°C for 10 min and 40 cycles of 95°C for 15 s followed by 60°C for 1 min. Undetermined or > 35 Ct values were replaced by 35. Renal transporters expression was then analyzed by the comparative $2^{-\Delta Ct}$ method with $\Delta Ct$ = Ct (target gene) – Ct (mean of the housekeeping genes finally retained). $2^{-\Delta Ct}$ were then normalized by log 2 transformation. Housekeeping genes finally retained for the $2^{-\Delta Ct}$ method (*NME4*, *CHFR* and *C16ORF62*) were selected using Genorm and Normfinder.

### 5. Statistical analyzes

The study had three objectives: (i) explore the impact of ischemia duration on the metabolomic content of perfusion liquids and on renal tubular transporters expression, (ii) explore relationships between metabolomic profiles and renal tubular transporters expression during HPM and (iii) find new biomarkers of IGF by comparing several parameters between patients with IGF or non-IGF. For the first objective, grafts were allocated to 1 out of 3 groups according to the perfusion duration in the machine: < 12h, between 12 and 20h, and > 20h. Metabolites intensities or concentrations (for those analysed with Cobas instrument) were normalized by the sum of all features and then log transformed. Auto scaling was performed prior to statistical analyses, in accordance with standard approaches for metabolomic analysis. A two-step statistical approach was performed for each objective. Univariate analyzes were performed using non-parametric tests, i.e. Wilcoxon and Kruskall-Wallis for two or more groups, respectively. P values were corrected for multiple statistical tests using the False Discovery Rate (FDR) method. Concerning multivariate exploration, unsupervised analysis by Principal Component Analyses (PCA) was performed prior to the use of different machine learning approaches (e.g. partial least-squares discriminant analysis (PLS-DA) and Random-Forest (RF)). Pathway analyzes were performed for metabolites identified as discriminant during multivariate analyzes. Correlation between normalized metabolites values and membrane transporters' expression (log2($2^{-\Delta Ct}$) was evaluated by Partial Least Squares regression (PLS). The MetaboAnalyst 5.0 computational platform (www.metaboanalyst.ca/faces/home.xhtml) was used for all the statistical analyzes except for the PLS analysis which was used to study the correlation between transporter expression and metabolite content, performed using the MixOmics package (version 1.6.3) in the R software (version 4.0.2).

**SDC, Figures:**

**Figure S1:** Pathway analysis based on metabolites only detected in perfusates.

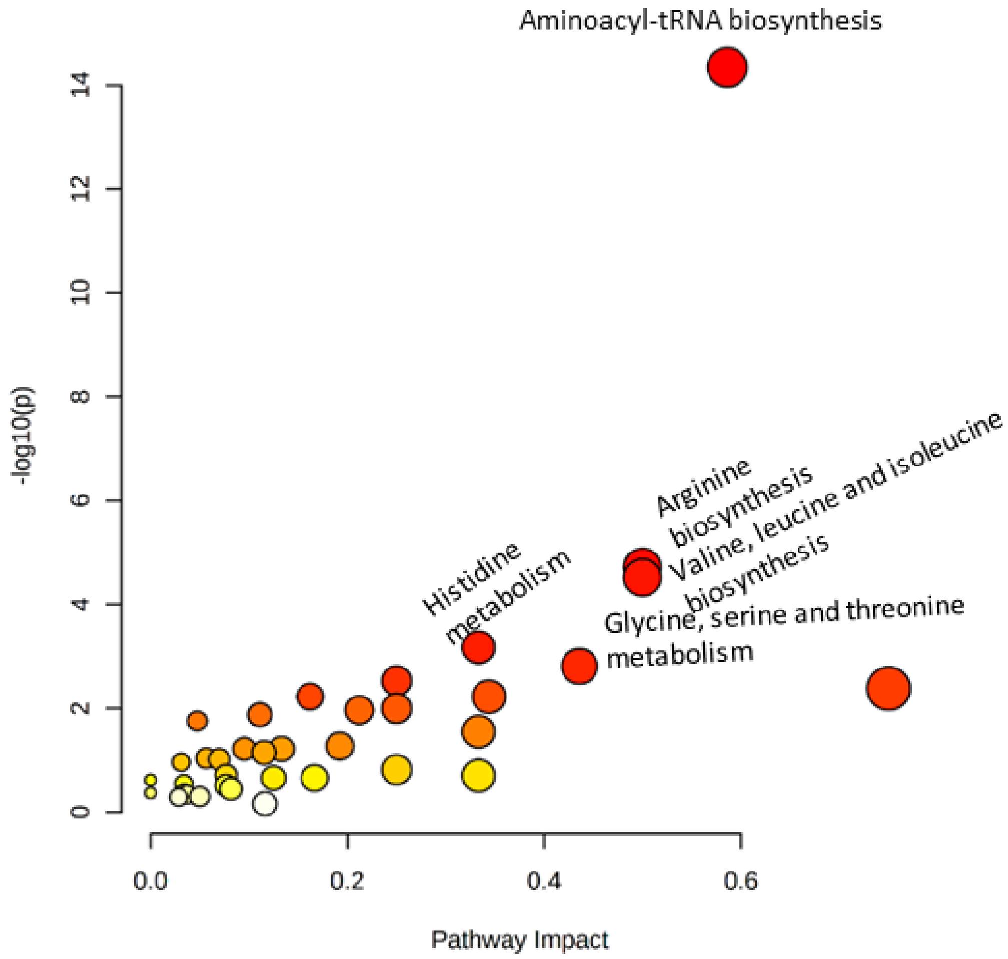

**Figure S2:** Correlation matrix between transporters found in pre-implantation biopsies. The dark red and dark blue colors indicate positive and negative correlations, respectively.

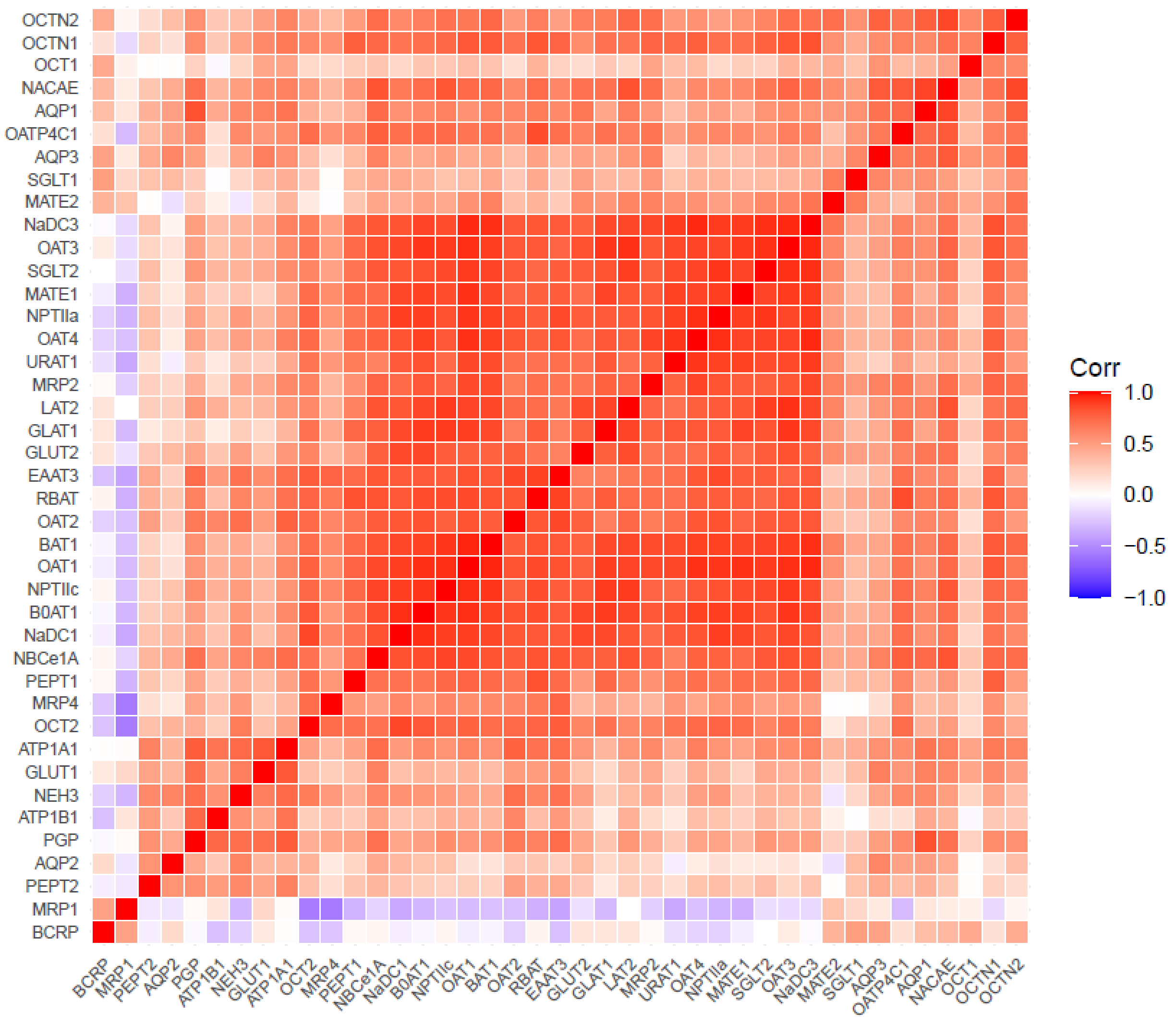

**Figure S3:** Dot plot of pathway enrichment analysis based on the most important metabolites, determined by Annova test, according to the perfusion time.

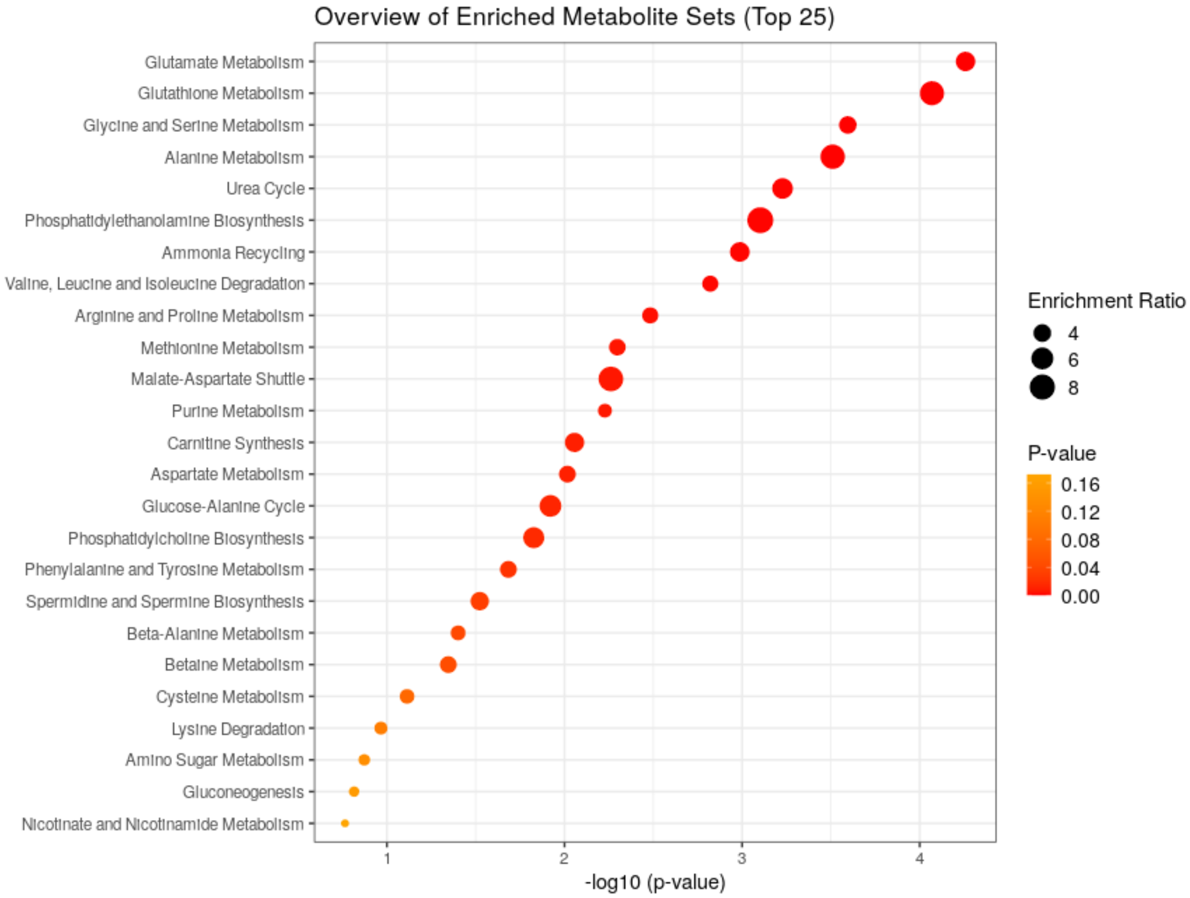

**SDC, Tables:**

**Table S1:** Custom-Designed TaqMan Low Density Array Card.

| Assay ID | Gene Symbol(s) | Amplicon Length |
|---|---|---|
| Hs00537914_m1 | *SLC22A6* | 68 |
| Hs00198527_m1 | *SLC22A7* | 69 |
| Hs00188599_m1 | *SLC22A8* | 144 |
| Hs01056646_m1 | *SLC22A8* | 76 |
| Hs00945829_m1 | *SLC22A11* | 82 |
| Hs00427552_m1 | *SLC22A1* | 79 |
| Hs01010726_m1 | *SLC22A2* | 70 |
| Hs00268200_m1 | *SLC22A4* | 76 |
| Hs00929869_m1 | *SLC22A5* | 65 |
| Hs00217320_m1 | *SLC47A1* | 74 |
| Hs99999905_m1 | *GAPDH* | 0 |
| Hs00945652_m1 | *SLC47A2* | 63 |
| Hs00960489_m1 | *ABCC2* | 62 |
| Hs00988720_g1 | *ABCC4* | 86 |
| Hs00988721_m1 | *ABCC4* | 141 |
| Hs01053790_m1 | *ABCG2* | 83 |
| Hs00184500_m1 | *ABCB1* | 67 |
| Hs01030727_m1 | *SLC22A12* | 64 |
| Hs00192639_m1 | *SLC15A1* | 76 |
| Hs01113665_m1 | *SLC15A2* | 69 |
| Hs00903842_m1 | *SLC9A3* | 77 |
| Hs00919316_g1 | *SLC13A2* | 72 |
| Hs00955744_m1 | *SLC13A3* | 68 |
| Hs01092910_m1 | *SLC34A1* | 84 |
| Hs02341453_g1 | *SLC34A3* | 94 |
| Hs00698884_m1 | *SLCO4C1* | 77 |
| Hs01573793_m1 | *SLC5A1* | 60 |

| | | |
|---|---|---|
| Hs00894642_m1 | *SLC5A2* | 75 |
| Hs00892681_m1 | *SLC2A1* | 76 |
| Hs01096908_m1 | *SLC2A2* | 65 |
| Hs00933601_m1 | *ATP1A1* | 76 |
| Hs00426868_g1 | *ATP1B1* | 89 |
| Hs01028916_m1 | *AQP1* | 96 |
| Hs00292214_s1 | *AQP2* | 87 |
| Hs00185020_m1 | *AQP3* | 63 |
| Hs01047033_m1 | *SLC4A4* | 68 |
| Hs01561483_m1 | *ABCC1* | 65 |
| Hs01384157_m1 | *SLC6A19* | 70 |
| Hs00909948_m1 | *SLC7A7* | 79 |
| Hs00794796_m1 | *SLC7A8* | 87 |
| Hs00374243_m1 | *SLC3A2* | 77 |
| Hs00942976_m1 | *SLC3A1* | 66 |
| Hs00204638_m1 | *SLC7A9* | 50 |
| Hs00188172_m1 | *SLC1A1* | 76 |
| Hs00943494_m1 | *CHFR* | 67 |
| Hs00359037_m1 | *NME4* | 70 |
| Hs01032748_g1 | *NASP* | 65 |
| Hs00220422_m1 | *C16ORF62* | 82 |

**Table S2:** MRM transitions, retention time and mass spectrometric conditions of metabolites

| Compound | Ionisation mode | Precursor ion | Product ion | | | | Retention time (min) |
|---|---|---|---|---|---|---|---|
| | | | 1 | 2 | 3 | 4 | |
| | | m/z | m/z | m/z | m/z | | |
| 2-Isopropylmalic acid | Negative | 175.15 | 113.05 | 115.05 | | | 5.2 |
| 1-Methylhistidine | Positive | 170.00 | 83.10 | 124.05 | | | 2.0 |
| 2-Aminoethanol | Positive | 62.15 | 27.10 | 44.10 | | | 1.8 |

| | | | | | | |
|---|---|---|---|---|---|---|
| 2-Ketoisovaleric acid | Negative | 115.20 | 43.20 | 59.20 | 71.05 | 4.8 |
| 3-Methyl-2-oxovaleric acid | Negative | 129.20 | 57.00 | 85.10 | 111.00 | 4.9 |
| 4-Aminobenzoic acid | Positive | 138.25 | 65.10 | 77.10 | | 5.3 |
| 4-Hydroxyproline | Positive | 132.10 | 68.05 | 86.05 | | 1.3 |
| 4-Pyridoxic acid | Positive | 184.00 | 148.10 | 166.10 | | 4.8 |
| 5-Glutamylcysteine | Positive | 251.10 | 84.10 | 122.10 | | 3.0 |
| 5'-Methylthioadenosine | Positive | 298.10 | 119.10 | 136.10 | | 6.2 |
| Adenine | Positive | 136.00 | 65.00 | 119.05 | | 4.9 |
| Adenosine | Positive | 268.10 | 119.00 | 136.05 | | 5.0 |
| Alanine | Positive | 89.90 | 44.10 | 45.30 | | 1.4 |
| alpha-keto-glutarate | Negative | 145.30 | 40.95 | 73.00 | 83.00 | 1.3 |
| Anthranilic acid | Positive | 138.00 | 65.05 | 120.10 | | 5.3 |
| Arginine | Positive | 175.20 | 60.10 | 70.10 | 116.00 | 1.8 |
| Aspartic acid | Positive | 134.00 | 70.10 | 74.05 | 88.10 | 1.3 |
| Biotin | Positive | 245.10 | 97.05 | 226.95 | | 6.0 |
| Choline | Positive | 104.10 | 45.10 | 60.05 | | 2.7 |
| Creatinine | Positive | 114.10 | 43.10 | 44.20 | 86.20 | 2.9 |
| Cystathionine | Positive | 223.00 | 88.05 | 134.00 | | 1.3 |
| Cystine | Positive | 241.00 | 73.90 | 119.95 | 151.95 | 1.3 |
| Deoxycytidine monophosphate | Positive | 308.10 | 95.10 | 112.05 | | 1.3 |
| D-Mannitol | Negative | 181.10 | 71.05 | 89.15 | 101.10 | 1.3 |
| D-Ribose | Negative | 149.30 | 59.05 | 71.15 | 89.05 | 1.3 |
| Gluconic acid | Negative | 195.20 | 74.90 | 99.00 | 129.05 | 1.2 |
| Glucosamine | Positive | 180.00 | 72.10 | 162.10 | | 2.0 |
| Glutamic acid | Positive | 147.90 | 56.10 | 84.10 | | 1.4 |
| Glutamine | Negative | 145.20 | 109.00 | 127.15 | | 1.4 |
| Glutathione | Positive | 308.00 | 130.10 | 179.10 | 276.00 | 3.2 |
| Glycine | Positive | 75.90 | 30.15 | | | 1.3 |
| Guanosine | Positive | 284.00 | 110.00 | 135.00 | 152.00 | 4.8 |
| Hexose (Glucose) | Negative | 179.20 | 59.05 | 89.10 | | 1.2 |
| Histidine | Positive | 155.90 | 56.10 | 82.70 | 110.10 | 1.7 |
| Hypoxanthine | Positive | 137.00 | 55.05 | 110.00 | | 3.1 |

| | | | | | | | |
|---|---|---|---|---|---|---|---|
| Inosine | Positive | 269.10 | 110.00 | 118.95 | 137.05 | | 4.8 |
| Isoleucine | Positive | 132.10 | 41.20 | 44.20 | 69.15 | 86.20 | 5.2 |
| Kynurenine | Positive | 209.10 | 94.10 | 118.10 | 145.90 | 192.05 | 5.9 |
| Lactic acid | Negative | 89.30 | 43.10 | 45.10 | 71.10 | 89.05 | 2.0 |
| L-Carnitine | Positive | 162.30 | 60.10 | 103.15 | | | 3.2 |
| Leucine | Positive | 132.10 | 30.05 | 43.35 | 86.05 | | 5.3 |
| Lysine | Positive | 147.20 | 56.10 | 84.20 | 130.10 | | 2.0 |
| Methionine | Positive | 149.90 | 56.10 | 104.10 | | | 3.2 |
| Methionine sulfoxide | Positive | 166.00 | 55.95 | 74.10 | 102.00 | | 1.4 |
| Niacinamide | Positive | 123.10 | 53.10 | 77.00 | 80.05 | | 3.5 |
| O-Phosphoethanolamine | Positive | 142.10 | 44.20 | | | | 1.2 |
| Ornithine | Positive | 133.10 | 70.10 | 116.05 | | | 1.6 |
| Oxidized glutathione | Negative | 611.10 | 143.05 | 306.00 | | | 4.8 |
| PAH | Positive | 195.00 | 65.20 | 92.20 | 120.15 | | 5.0 |
| Phenylalanine | Positive | 166.10 | 77.10 | 103.10 | 120.10 | | 5.7 |
| Pipecolic acid | Positive | 130.10 | 56.10 | 84.05 | 112.10 | | 2.6 |
| Proline | Positive | 116.10 | 28.05 | 43.20 | 70.15 | | 1.7 |
| Riboflavin | Positive | 377.00 | 172.00 | 197.00 | 243.05 | | 5.2 |
| Serine | Positive | 105.90 | 42.30 | 60.10 | 70.10 | | 1.3 |
| Taurine | Negative | 124.35 | 64.10 | 79.95 | | | 1.2 |
| Threonine | Positive | 120.10 | 56.05 | 74.15 | 102.10 | | 1.3 |
| Thymidine | Positive | 243.10 | 109.00 | 116.00 | 127.10 | | 4.8 |
| Thymine | Positive | 127.10 | 54.05 | 110.05 | | | 4.8 |
| Tryptophan | Positive | 205.10 | 91.00 | 118.00 | 146.10 | 188.15 | 6.8 |
| Tyrosine | Positive | 182.10 | 77.10 | 91.10 | 136.10 | | 4.9 |
| Uracil | Positive | 113.00 | 70.00 | | | | 2.1 |
| Uric acid | Negative | 167.10 | 96.20 | 123.95 | | | 2.4 |
| Uridine | Positive | 245.00 | 70.00 | 96.00 | 113.05 | | 3.2 |
| Valine | Positive | 118.00 | 55.10 | 57.10 | | | 2.9 |
| Xanthine | Negative | 151.00 | 42.00 | 108.00 | | | 3.1 |
| Xanthosine | Positive | 284.90 | 135.95 | 153.05 | | | 4.8 |
